# The impact of pre-existing eye donation awareness of the next of kin on donation rate after grief counseling - A cross-sectional study

**DOI:** 10.1101/2023.11.15.23298568

**Authors:** Oshin Puri, Neeti Gupta, Sanjeev Mittal

## Abstract

This cross-sectional study was conducted to assess the impact of pre-existing eye donation awareness of the next of kin on the donation rate after grief counseling with due ethical approval. The N=164 most stable next-of-kin, mostly the Brother (20.7%) or Father (20.1%) of the deceased, were approached by the Eye Donation Counselor (EDC). After assessing their awareness about eye donation through the “*Awareness and Perception on Eye Donation*” questionnaire (Ronanki, V.R, et. al), the EDC grief counseled the next of kin. 84.8% of all participants were aware of eye donation. The mean awareness, knowledge, and perception scores of the study population were 2.46 +/- 1.34 out of 4, 2.73 +/- 2.37 out of 6, and 1.79 +/- 1.72 out of 4 respectively. Eye care professionals (N=105 (64%)) and mass media (N=61 (37.2%)) were identified as the most common sources of information. While 52.4% expressed willingness to donate, only 7.3% donated and there was 1 voluntary donation. Counseling and the belief of eye donation being a noble deed were identified as the major motivators, and objections by other family members, and religious beliefs were the major barriers. There is no significant association between eye donation and the pre-existing awareness of the next of kin regarding eye donation. Although awareness is associated with the increased willingness to donate eyes.

## Introduction

The International Agency for the Prevention of Blindness (IAPB) reports that a total of 1.1 billion people across the globe have vision loss, of which 43 million are blind. (1) The 2018 Global Causes of Vision Loss Estimates from IAPB reported that 2.4% of global visual impairment/blindness results from corneal causes amounting to a total of 6.17 million people worldwide. (2, 3) The Eye Banking Association of India (EBAI) reports that about 1.1 million Indians suffer from corneal blindness. (4) Corneal blindness might result from degeneration, dystrophy, infection, inflammation, and secondary damage and is the most common cause of reversible blindness. (5,6)

With limited treatment alternatives, corneal transplantation has emerged as the definitive treatment for enhancing visual acuity and quality of life. (5, 7, 8) Despite the success of corneal transplants, only 25% of the 1,00,000 transplants required in India are being conducted annually. (4) Scarcity of donor corneas has been identified as the major cause of the same. (9) Strategies like Community Awareness Interventions (CAI), Eye Donation Pledges, Grief Counseling, and the Hospital Cornea Retrieval Program (HCRP) are being encouraged to curb the scarcity. (10, 11)

CAIs enhance donor willingness and registry signing. (12, 13) Limited evidence is available evaluating actual donation post-CAIs. (12, 13) Willingness and registry signing might be suitable measures to evaluate conversion rates for live tissue/organ donations. But post-death, when the next of kin holds the right to donate (irrespective of the deceased’s pledge), the willingness of the deceased might be a deceptive outcome measure. (14) This is especially important since the individual’s wish to donate has been shown to have no effect on the family’s decision to donate. (14)

CAIs targeting families rather than individuals might have better outcomes, but evidence suggests that pre-existing awareness as a whole does not contribute to enhancing donation. (11, 14-17) Thus, the impact of such resource-intensive CAIs continues to remain debatable. (14-16, 18, 19) The need to enhance donation, the uncertain impact of CAIs, and the prevalence of misinformation have led to grief counseling emerging as the mainstay to enhance eye donation. (15, 17, 20)

An Eye Donation Counsellor (EDC) is a science graduate (Bachelor of Science or equivalent) having undergone a 3-month training in grief counseling and cornea retrieval from an eye donation training center. Through HCRP, EDCs at the eye bank are notified of every death in the hospital. The EDC then visits the deceased, identifies the most stable next of kin, and approaches them for grief counseling. Since cornea must be harvested within 6-8 hours post-death, grief counseling is initiated as soon as possible after death. Grief counseling usually starts with empathizing with the next of kin, and assessment of their awareness, followed by a discussion regarding the need for donation, doubt addressal, and consent. Grief counseling has been shown to significantly enhance eye donation. (15, 17, 20) The evident advantage grief counseling has over CAIs is that it is directed toward the primary stakeholders of donation: the next of kin, and is delivered just before donation consent is taken.

There is a paucity of literature methodically assessing the impact of awareness on actual eye donation. Thus, the primary objective of this study is to assess the impact of pre-existing eye donation awareness of the next of kin on the donation rate after grief counseling.

## Material & Methods

### Study Design

This study was conducted as a cross-sectional study.

### Consent and Ethical Considerations

A well-informed consent was taken from all participants. The study was conducted with due ethical clearance from the Institutional Ethics Committee (IEC) of the All India Institute of Medical Sciences (AIIMS), Rishikesh, India received via letter number 262/IEC/STS/2022.

### Study Population

The most stable next of kin of patients deceased in the critical care areas of the hospital and those brought to the mortuary were invited to participate in the study. The most stable individual was approached since donation in India is an opt-in system, necessitating counseling for each death.

### Sample Size

The sample size for this cross-sectional study was calculated by -

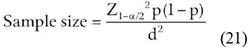

Where,

Z_1-a/2_ = Is standard normal variate. Considering a 5% type 1 error (P<0.05) it is 1.96.

p = Expected proportion in population based on previous studies or pilot studies.

d = absolute error of precision.

The proportion of corneas donated and deaths in the institute (p) was considered to be 8% based on the eye bank’s records. For a type-1 error of 5% and a 5% absolute error of precision, the minimum number of participants that were to be enrolled in the study was calculated to be **113**. Data from **164** participants were collected.

### Study Tools

#### Eye Donation Awareness

The pre-existing awareness of the most stable next of kin regarding eye donation was assessed through “*Awareness and Perception on Eye Donation*” an open-access 15-item questionnaire developed by Ronanki, V.R. et. al. used with due permission from the corresponding author. (22) The questionnaire has been tested for its face, content, and flow validity as a tool to assess awareness, sources of information, knowledge, perception, barriers, enablers, and willingness to donate among Indian patient attendants.

#### Grief Counseling

The next of kin was counseled for eye donation following the institute’s Grief Counseling Standard Operating Protocol (SOP) under HCRP in use since the inception of the eye bank (2019).

#### Donation

The individual’s willingness to donate the deceased’s eyes was then assessed. Data regarding actual donations after grief counseling and voluntary donations were collected from the Eye Bank HRCP records.

### Methodology

The most stable individual next of kin was identified by the EDC. The relation of this next of kin to the deceased and their pre-existing awareness regarding Eye Donation was assessed through an interview (based on “*Awareness and Perception on Eye Donation’’*). Once the next of kin was counseled, their willingness to donate the deceased eyes was assessed along with the reasons underlying the same. Whether the family actually donated was later recorded. Data regarding voluntary donation was also recorded.

### Outcome Measures

This study generated data in the forms of -

1. Categorical variables of an individual being aware/unaware of eye donation, willingness to donate, and actual donation.
2. Discrete interval variables quantifying the (extent of) awareness, knowledge, and perception of the study population regarding eye donation.
3. Description of common sources of information, barriers, and enablers of donation, and the general population’s suggestions to enhance eye donation.

### Statistical analysis

The primary objective of the study was to assess the association between eye donation and next to the kin’s pre-existing awareness. This association was tested by the Chi-Square test. Considering a probability level of 0.05 at a degree of freedom = 1, Chi-Square greater than the Chi-Square Statistic (here, 3.841) was considered to imply a significant association between the variables. The study also assesses the association between the next of kin’s willingness to donate and his/her awareness, considering Chi-square > 3.84 to imply a significant association.

Secondary objectives of generating preliminary data regarding awareness, knowledge, and perception regarding eye donation, sources of information, barriers, and enablers, willingness to donate, and actual donations have been descriptively reported.

## Observations and Results

n=54 (32.9%) participants identified as females. Other demographics could not be recorded as discomfort was identified among participants in discussing their own age, educational qualifications, occupation, and religion in the given circumstances. The relation of the participants to the deceased (Figure-1) suggests that the brother (20.7%) or the father (20.1%) were most commonly identified as the most stable next of kin.

**Figure-1:**
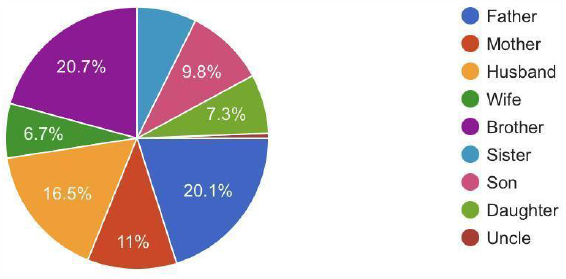
Relation of the study participant (most stable next of kin) to the deceased.

Assessment of awareness suggested that N=139 (84.8%) study participants were aware of (had heard of) eye donation. N=89 (45.7%) knew about the nearby eye bank. N=119 (72.6%) knew that eye donation was a post-death donation and N=71 (43.3%) knew that prior pledging of eyes wasn’t necessary to donate eyes. The cumulative awareness score of all the participants was 2.46 +/- 1.34 of a maximum of 4. (Table-1)

**Table-1:** Variable description of Awareness, Knowledge, Perception, and Cumulative Awareness and Perception of the study population regarding eye donation.

|  | Max | Mean | SD | 25th Percentile | Median | 75th Percentile | IQR |
| --- | --- | --- | --- | --- | --- | --- | --- |
| **Awareness** | 4 | 2.46 | 1.34 | 2 | 3 | 4 | 2 |
| **Knowledge** | 6 | 2.73 | 2.37 | 1 | 1.5 | 5 | 4 |
| **Perception** | 4 | 1.79 | 1.72 | 0 | 1 | 4 | 4 |
| **Cumulative Awareness and Perception** | 14 | 6.99 | 5.12 | 3 | 5 | 13 | 10 |

Assessment of knowledge suggested that N=66 (40.2%) study participants were aware of the definition of eye donation. N=44 (26.8%) knew the window period for eye donation, N=76 (46.3%) knew that only cornea is retrieved in eye donation, and N=67 (40.9%) knew that donated corneas are used to replace the cornea of another individual’s eye(s). N=125 (76.2%) knew that donated eyes could give eyesight to 2 blind individuals, and N=75 (43.3%) knew eye donation does not disfigure the donor’s face. The cumulative knowledge score of all the participants was 2.73 +/- 2.37 of a maximum of 6. (Table-1) Eye care professionals namely ophthalmology consultants, residents, and nurses, and optometrists were the most common sources of information regarding eye donation followed by social media. (Figure-3)

Assessment of the perception regarding eligibility to donate suggested that N=72 (43.9%) knew that there is no age limit to donate eyes. N=94 (57.3%) knew that donation was not limited to any gender, N=67 (40.8%) knew that individuals with prior use of spectacles could also donate eyes and N=61 (37.2%) knew individuals having a history of chronic illness could also donate eye (Figure-2). The cumulative perception score of all the participants was 1.79 +/- 1.72 out of a maximum of 4. (Table-1)

**Figure-2:**
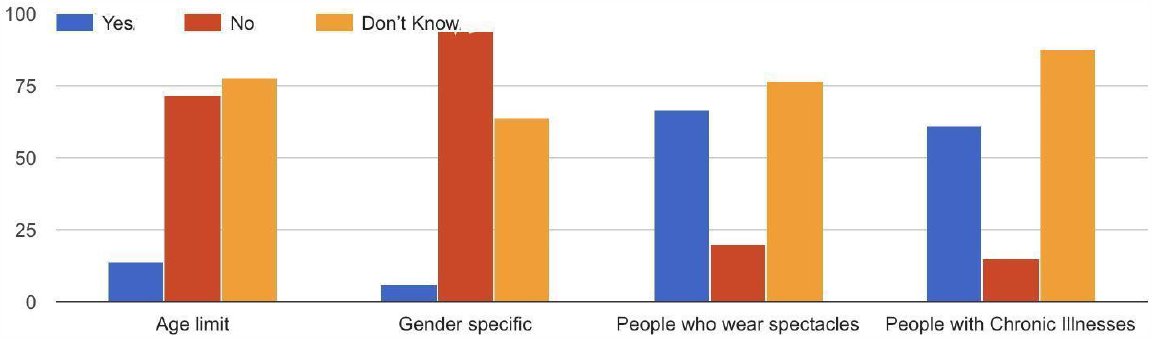
Bar-graphs representing the proportion of study participants aware of the eligibility to donate eyes not being limited to (a) any age group, (b) any gender group, (c) no prior use of spectacles, and (d) absence of chronic illnesses.

**Figure-3:**
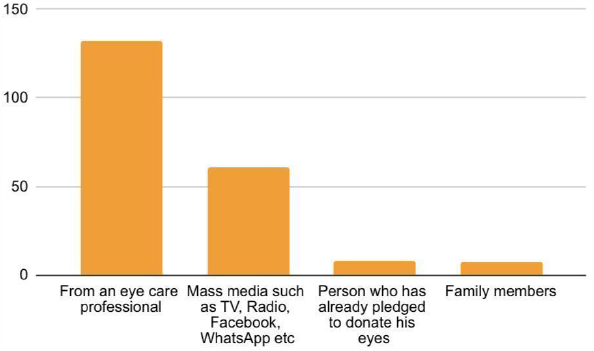
Sources of Information regarding eye donation.

A total of N=86 (52.4%) participants expressed willingness to donate the deceased eyes but only N=12 (7.31%) donations were made. During the same period, the eye bank received N=1 voluntary eye donation. Assessment of the motivators and barriers to willingness to donate suggested that the strongest motivator was grief counseling (encouraging N=62 (37.8%) participants) followed by eye donation being a noble work (encouraging N=59 (35.98%) participants). (Figure-4a) Objection by other family members (N=105, 64.02%) and the lack of awareness about eye donation (N=61, 37.19%) were identified as the major barriers to donation. (Figure-4b)

**Figure-4:**
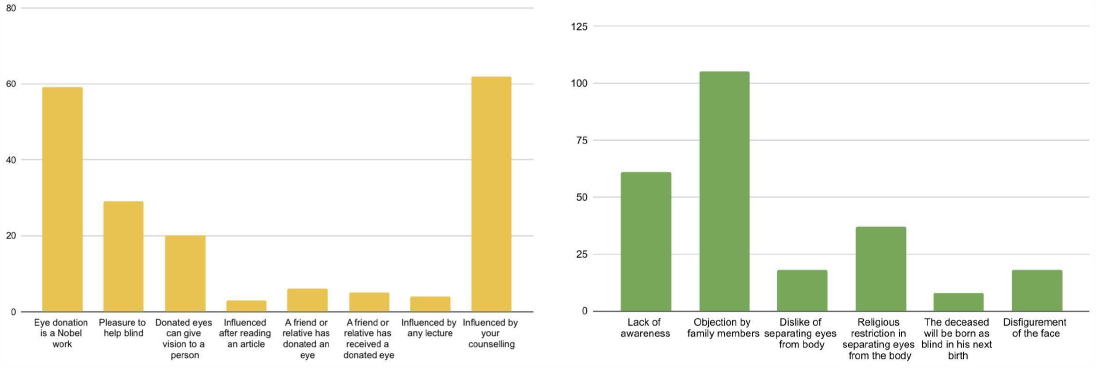
Bar graphs representing (a) Motivators/Enables (b) Barriers to eye donation.

### Chi-Square test

Chi-Square for the association between eye donation and pre-existing awareness of the next of kin was 0.95 < Chi-Square Statistic (3.841), implying p > 0.05 suggesting that there is no significant association between eye donation and pre-existing awareness of the next of kin.

Chi-Square for association willingness to donate eyes and pre-existing awareness of the next of kin was > 10 > Chi-Square Statistic (3.841), implying p < 0.05 suggestive that there is a significant association between willingness to donate eyes and pre-existing awareness of the next-of-kin.

## Discussion

Brothers and the Fathers of the deceased were most frequently identified as the most stable next of kin. Similar results have also been reported in the past where 41.8% of the most stable next of kin were siblings. (20)

The current study reported 84.8% awareness, similar to that reported in other parts of North India. (14) The existing literature also reports differences in awareness about eye donation within the same region suggesting that results regarding awareness can vary depending on sampling techniques, and period of assessment. (14, 22-26) In our study, 85% awareness might be due to the sampling of attendants of chronically ill patients regularly visiting the hospital, hence more likely to attend a CAI.

N=119 (72.6%) of the population is aware of eye donation being a post-death donation, and only N=71 (43.3%) aware of prior pledging not being necessary for donation. These numbers were definitely lower than similar studies where 80.3% and 81.1% of eye donation stakeholders from Srikakulam and 98.1% and 78% of adults from rural Pondicherry were aware of the occasion of donation and prior pledging. (22, 23, 25). N=10 (6.1%) believed that eye donation is a live donation, compromising donor vision, N=37 (22.6%) thought that eye donation disfigures the face and N=39 (23.8%) had concerns regarding the use of the donated tissue, which has also been reported as a barrier to eye donation in the past. (20)

Addressing these misconceptions is imperative to enhance donations. Grief counseling addresses these doubts and myths just before consent and enhances donation. But, such misinformation also needs to be addressed in the community. Only one voluntary donation during the study period emphasizes the need for the latter. But, voluntary donation requires a larger group to agree, that too without any professional counseling, thus CAIs need to be family-centered.

Regarding eligibility to donate, most participants weren’t aware of any restrictions to eye donation, notably different from a study from South India where the awareness was much higher. (22) N=89 (45.7%) participants were aware of the nearest eye bank, similar to the published literature. (22, 23, 25) Only N=44 (26.8%) were aware of the time until corneas can be donated. It is noteworthy, that even those aware of eye donation, might not know the eligibility to donate, the technicalities of pledging, or the nearest eye bank.

This technical & logistic information is specifically important for voluntary donation. This suggests that CAIs must be carefully curated to not just motivate donation but also deliver necessary logistic information free from medical jargon. The same is reiterated by the highly dispersed cumulative knowledge and perception scores. While a few had a thorough knowledge of the technicalities of eye donation, the majority had a limited understanding of the same.

The study also identified eye care professionals (namely ophthalmology consultants, residents, and nurses, and optometrists) and mass media as the most common sources of eye donation-related information. Existing literature also reports mass media to be a major source of information. (25)

While N=86 (52.4%) participants expressed willingness to donate, only N=12 (7.31%) donations were made. Gogate B. et. al. in their letter to the Indian Journal of Ophthalmology discussed this disparity and documented that while the willingness to donate eyes has always been reported to be high, the actual donations have been extremely low. (27) The results of the Chi-square test revealed that while awareness was associated with the “willingness to donate”, it had no association with actual donations. The findings of the current study are in line with the existing evidence suggesting that while CAIs and increased awareness, may contribute towards increasing “willingness to donate”, it may not impact actual donations, (11, 15, 19, 24, 27) especially among those grief counseled post-death. (14, 25) Literature assessing the same in tissue/organ donations other than eye also reported similar results. (12, 13) Gogate B. et. al. in their attempt to explain the aforementioned identified that while willingness to donate stems from a belief in doing something “good” the same belief is difficult to maintain in traumatic situations of the loss of a loved one. (27)

The major motivators to donation were grief counseling, doing a noble deed, pleasure in helping a blind individual, and the belief that donated eyes can give eyesight to a blind individual. Several studies in the past have also reported similar conclusions. (11, 14, 15) It is also worth noting that emotions and beliefs are the second most important motivator and thus must be respected during grief counseling. This study also suggests that while Grief Counseling is an effective method of enhancing eye donations, its impact can be further enhanced by reiterating how noble eye donation is and how it may help a blind individual see the world.

The current study identified objections by family members other than the one counseled to be the most common barriers to donation, similar to the existing literature. (25, 28) Addressing this barrier raises the dilemma if grief counseling should also need to be family-centered. While counseling 2-3 individuals instead of one might seem a suitable alternative, practically identifying and communicating with even one stable next of kin within hours of death is challenging. Furthermore, members of the family looking into the discharge and distant relatives residing close by might arrive after counseling. Such influences might be difficult to predict or address, further raising concerns as to how objections by accompanying family members can be addressed.

Religious beliefs and the belief of the deceased being born blind in the next life emerged as the next most common reasons behind the denial to donate. Religious beliefs have been reported as a major determinant of willingness to donate on multiple occasions in the past as well. (24, 25, 29) Religion might be the most delicate barrier to address, although the “Catalyst” approach suggested by Gogate et. al. might help. (27) They suggest involving persons of faith since they have significant influence and the potential of enhancing donation. (27)

A case-control study design between donors and non-doners without grief counseling might give more reliable information regarding the impact of awareness on donation. But, this might compromise donation since potential donors who might have donated after counseling would not have been approached. Thus, the current study was conducted to generate maximum data with minimum donation compromise. Awareness being analyzed as a categorical variable might be another limitation of the current study.

## Conclusion

The current study reports 7.31% of donations after grief counseling. There is no significant association between eye donation and the pre-existing awareness of the next of kin. Although awareness is associated with willingness to donate eyes, but does not necessarily translate to consent and retrieval of eye tissue. Major motivators identified in the study, are grief counseling, nobility of eye donation, and its benefits to the recipient.

## Data Availability

All data produced in the present study are available upon reasonable request to the authors.

